# A Label-free Optical Biosensor-Based Point-of-Care Test for the Rapid Detection of Monkeypox Virus

**DOI:** 10.1101/2024.07.03.24309903

**Authors:** Mete Aslan, Elif Seymour, Howard Brickner, Alex E. Clark, Iris Celebi, Michael B. Townsend, Panayampalli S. Satheshkumar, Megan Riley, Aaron F. Carlin, M. Selim Ünlü, Partha Ray

**Author notes:** Address correspondence to: Partha Ray, M. Selim Ünlü.

## Abstract

Diagnostic approaches that combine the high sensitivity and specificity of laboratory-based digital detection with the ease of use and affordability of point-of-care (POC) technologies could revolutionize disease diagnostics. This is especially true in infectious disease diagnostics, where rapid and accurate pathogen detection is critical to curbing the spread of disease. We have pioneered an innovative label-free digital detection platform that utilizes Interferometric Reflectance Imaging Sensor (IRIS) technology. IRIS leverages light interference from an optically transparent thin film, eliminating the need for complex optical resonances to enhance the signal by harnessing light interference and the power of signal averaging in shot-noise-limited operation to achieve virtually unlimited sensitivity. In our latest work, we have further improved our previous ‘Single-Particle’ IRIS (SP-IRIS) technology by allowing the construction of the optical signature of target nanoparticles (whole virus) from a single image. This new platform, ‘Pixel-Diversity’ IRIS (PD-IRIS), eliminated the need for z-scan acquisition, required in SP-IRIS, a time-consuming and expensive process, and made our technology more applicable to POC settings. Using PD-IRIS, we quantitatively detected the Monkeypox virus (MPXV), the etiological agent for Monkeypox (Mpox) infection. MPXV was captured by anti-A29 monoclonal antibody (mAb 69-126-3) on Protein G spots on the sensor chips and were detected at a limit-of-detection (LOD) - of 200 PFU/ml (∼3.3 attomolar). PD-IRIS was superior to the laboratory-based ELISA (LOD - 1800 PFU/mL) used as a comparator. The specificity of PD-IRIS in MPXV detection was demonstrated using Herpes simplex virus, type 1 (HSV-1), and Cowpox virus (CPXV). This work establishes the effectiveness of PD-IRIS and opens possibilities for its advancement in clinical diagnostics of Mpox at POC. Moreover, PD-IRIS is a modular technology that can be adapted for the multiplex detection of pathogens for which high-affinity ligands are available that can bind their surface antigens to capture them on the sensor surface.

## INTRODUCTION

Monkeypox virus (MPXV) is an enveloped double-stranded DNA virus belonging to the Orthopoxvirus genus in the *Poxviridae* family (Sklenovská and Van Ranst, 2018). MPXV causes Monkeypox (Mpox), an infectious zoonotic disease first reported in 1958 (von Magnus et al., 1959). In the past, Mpox was considered a rare sporadic disease with a limited capacity to spread among humans (Organization and others, 1984). However, the recent rapid global spread of this infection, which is now recognized as the most critical orthopoxvirus infection in humans in the post-smallpox eradication era, has brought this neglected disease back into the spotlight.

Based on their genome sequence, MPXV has two major types: clade I and II (Likos et al., 2005). The clade I virus, endemic to central Africa, is particularly virulent, with human case fatality rates during some outbreaks estimated to be around 10%. A current Mpox (Clade I) outbreak in the Democratic Republic of Congo (DRC) is a testament to this. The country has reported the most significant surge of Mpox cases ever recorded, with over 20,000 suspected cases and more than 1000 deaths since January 2023 (CDC, 2024a). The Clade II virus, formally known as the West African clade, can be further categorized into two phylogenetically distinct subclades: Clade IIa and IIb. Clade II results in less severe infection than Clade I; however, the global Mpox outbreak that started in 2022, caused by Clade IIb, serves as a stark reminder of the potential worldwide impact of these viruses. The rapid spread of Mpox initially led the WHO to declare a Public Health Emergency of International Concern (PHEIC) on July 23, 2022, which was later lifted on May 10, 2023, following the recommendation from the International Health Regulations (IHR) Emergency Committee noting that progress was made in managing the disease outbreak. However, it should be noted that Mpox cases are still being detected in non-endemic countries, and as of May 2024, 95,912 confirmed cases in over 118 countries had been reported, resulting in 184 deaths (CDC, 2024b).

MPXV is transmitted primarily through close contact with infected wild animals or individuals and direct contact with body fluids (CDC, 2024c). There is currently no treatment approved by the U.S. Food and Drug Administration (FDA) for Mpox. However, a clinical trial is underway to test the efficacy of Tecovirimat (an antiviral drug against smallpox infection) in treating Mpox (Stomp, 2024). A smallpox vaccine (JYNNEOS™) has also been recently approved in the USA by the FDA under Emergency Use Authorization (EUA) for the prevention of Mpox infection in adults (FDA, 2022); however, due to the limited supply of the vaccine, its equitable distribution and access is a significant concern (Kahn et al., 2023).

Therefore, the most effective measure to prevent the further spread of this infection is to make a timely diagnosis of the MPXV and isolate infected individuals. MPox’s clinical manifestations, such as skin lesions, fever, headache, muscle soreness, and lymphadenopathy (swollen lymph nodes), are often atypical and can be easily mistaken for other prevalent infections (Hussain et al., 2022). Thus, diagnostics based on clinical criteria alone are challenging and underscore the need for molecular-based diagnosis, which can assist physicians in managing the disease and help health authorities implement effective countermeasures (Bourner et al., 2024).

Confirmatory tests of Mpox infection are performed by quantitative real-time PCR (qPCR) on the isolated virus DNA collected from the patient’s lesion specimens. However, the large central region of the Orthopoxvirus genus is highly conserved. Therefore, choosing proper target regions on the Mpox genome for PCR is essential to prevent cross-reactivity of the tests and differentiate between other orthopoxvirus species. Also, deletion in the MPXV genome results in false negative results in the PCR detection methods (Garrigues et al., 2022). Moreover, PCR-based tests, while accurate, can only be conducted in dedicated laboratories, making them unsuitable for rapid diagnostic tests at the point of care (POC). Few recent studies based on other nucleic acid amplification and detection methods, e.g., Loop-Mediation Isothermal Amplification (LAMP), Recombinase Polymerase Amplification (RPA), and Clustered Regularly Interspaced Short Palindromic Repeats (CRISPR)-Cas system, have demonstrated the Mpox diagnostic capability at the POC (Wang et al., 2024; Yu et al., 2023). However, factors like pre-processing steps for nucleic acid extraction from specimens, complicated designing of multiple primers for the viral genome amplification, high costs of reagents, and non-specific amplification producing false-positive results are some of the technical and logistical drawbacks that need to be addressed before their deployment at POC.

Antigen-antibody-based POC assays like the Lateral Flow Immuno Assay (LFIA) are affordable, can be conducted at home or outpatient clinics by untrained personnel, and are thus ideal for rapid testing of viruses (Ince and Sezgintürk, 2022). However, compared to PCR-based assays, they often lack sensitivity, specificity, and quantitative readout to detect viral antigens (Posthuma-Trumpie et al., 2009). There is no FDA-approved commercial antigen-based assay for detecting Mpox (CDC, 2024d). The serological assay that detects antibodies developed in patients in response to Mpox infection can also be used for retrospective analysis at the point of care. However, the kinetics of antibody production in response to infection varies in patients, especially in immunocompromised patients like people living with AIDS. Additionally, these tests are not specific for Mpox, often resulting in false positive results due to the patients’ prior smallpox vaccination history (Matusali et al., 2023).

These limitations of currently available nucleic acid or antigen-based tests highlight the urgent need to develop better point-of-care diagnostic tests for Mpox. An ideal diagnostic test would combine the sensitivity and specificity of quantitative nucleic acid-based tests with the user-friendly nature of rapid antigen-based point-of-care tests. This is particularly crucial in low-resource settings, where untrained personnel can utilize a simple diagnostic system that does not require expensive equipment to provide rapid, reliable, and affordable test results.

Optical sensors have been widely applied to detect viruses due to their simple use, cost-effectiveness, and direct detection capability (Maddali et al., 2021). An optical biosensor can detect viral nucleic acids, antigens, whole viruses, or antibodies produced in response to the viral infection for viral diagnostics. Some examples of these sensors include fluorescence-based optical sensors, surface plasmon resonance (SPR), optical resonators, and interferometry-based methods. Fluorescence-based sensors utilize fluorophore-attached detection molecules to bind to the viral target molecules captured on the surface via a virus-specific capture probe. Although fluorescence-based systems offer multiplexed detection and simpler workflows compared to lab-based tests such as ELISA, issues such as limited sensitivity, photobleaching, and non-specific binding affect the performance of these sensors (Seymour et al., 2023a). Label-free biosensors like SPR and optical resonator-based platforms are promising for POC viral diagnostics and can offer simple, direct, and sensitive detection (Baaske and Vollmer, 2012; Pandey et al., 2022). However, despite being good candidates for POC diagnostic devices, these platforms have certain limitations. SPR sensors suffer from the bulk effect (background signal due to buffer and temperature changes), low selectivity, limited multiplexing, and high substrate costs (Hassan et al., 2021; Nguyen et al., 2015). Optical resonator-based sensors are sensitive to temperature changes, and their substrates require complicated fabrication processes (Rho et al., 2020).

In contrast to these techniques, interferometry-based sensors gained attention owing to their simple and robust transduction mechanisms and minimal dependence on external factors. One such interferometric platform is a single-particle interferometric reflectance imaging sensor (SP-IRIS) composed of an LED-based imaging system and a layered silicon/silicon dioxide microarray chip (Daaboul et al., 2010). SP-IRIS can visualize single nanoparticles (NPs), both synthetic (e.g., polystyrene, gold NPs) and natural (e.g., intact viruses, extracellular vesicles), captured on the sensor surface via immobilized capture probes (Daaboul et al., 2016; George G Daaboul et al., 2014; Scherr et al., 2016). Digital detection by SP-IRIS is achieved by collecting all the light emanating from the sensor surface in the presence of virus particles. The corresponding signal correlates to the particle’s polarizability (or size). Discerning the optical signatures becomes particularly challenging as the size gets smaller. Therefore, SP-IRIS utilizes z-scan acquisition (a stack of images taken from different focal positions) to capture the defocus signature unique to sub-diffraction limited scattering object (viruses on sensor surface). However, z-scan acquisition imposes two major drawbacks: (i) acquiring z-stacks requires repeatable and high-resolution scanning optics. (ii) the computationally expensive algorithms are required to process the z-stacks.

In this work, we present the first demonstration of multi-spectral Pixel Diversity IRIS (PD-IRIS) for label-free and rapid detection of MPXV, where the necessary optical signature is encoded into multiple wavelengths. The major improvement introduced in PD-IRIS is that target particles can be detected at a single snapshot, making the precise z-scanning parts obsolete. This is achieved by exciting the particles under multi-parametric light and collecting the resulting waves on a CMOS array decorated with a filter array. The term pixel-diversity stems from this multi-parametric imaging technique. We utilize a monoclonal antibody (mAb 69-126-3), developed by the Centers for Disease Control and Prevention (CDC) (Roumillat et al., 1984), to capture intact MPXV on the PD-IRIS sensor chip surface spotted with Protein G (Figure 1). The antibody binds to the A29 protein with high affinity and specificity (Laura J Hughes et al., 2014). MPXV A29L is a homolog of vaccinia virus A27 associated with a mature virus membrane that binds to the host cell surface heparan sulfate and is essential for membrane fusion (Chang et al., 2013). We demonstrate that the new PD-IRIS prototype can rapidly detect MPXV (sample-to-result within 20 minutes) with higher sensitivity than the traditional laboratory-based Enzyme-Linked Immunosorbent Assay (ELISA) method. The setup can also be reconfigured in a compact manner to fit in a table-top device, allowing easy and quick result interpretation; thus, our assay, independent of any cell culture or sample pre-processing before testing like in SP-IRIS (Monroe et al., 2013), can be performed by untrained personnel and adapted to detect Mpox infection at the POC. Thus, PD-IRIS enables the practical implementation of digital virus detection, allowing for an automated, compact, and robust POC configuration.

**Figure 1:**
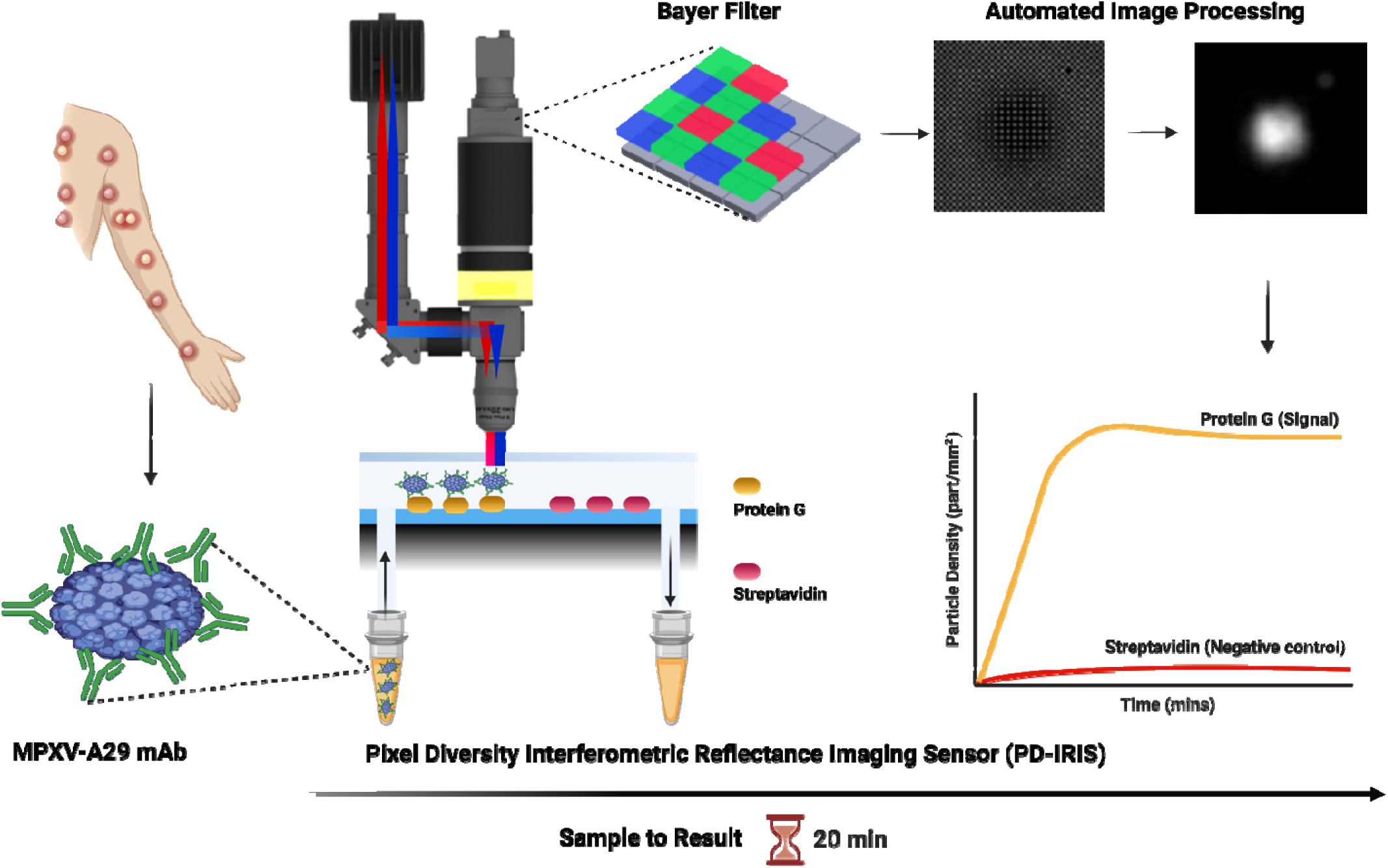
A schematic showing PD-IRIS workflow for MPXV detection in a POC format. First, the sample (MPXV) is mixed with specific detection antibodies (A29 mAb), and then, this mixture flows over the PD-IRIS chip assembled in a microfluidic cartridge. As the antibody-decorated viruses are captured on the surface-spotted protein G, bound particles appear as white dots on the camera following an image processing step. The overall signal is calculated as particle density (particles/mm^2^), and the binding curves are generated to show the signal on protein G and negative (streptavidin) spots. The figure was made using the *BioRender* software.

## MATERIALS AND METHODS

### Viral growth, titration, and inactivation

MPXV was isolated in 2022 from a Mpox-positive patient sample on VeroE6 cells (ATCC, CRL-1586). Serial dilutions of the patient sample were made in DMEM (Corning, Catalog number 10-013CM) + 2% FBS (Biowest, Catalog number S1520) + 1x Antibiotic/Antimycotic (Thermo Fisher Scientific, Catalog number: 15240062) + 10mM HEPES (Thermo Fisher Scientific, Catalog number: 15630080) and applied to cells in the same medium but with 10% FBS. Cells were scraped upon the appearance of the Cytopathic Effect (CPE). The virus was confirmed to be Clade II by amplification and Sanger sequencing of two diagnostic regions (positions 46,239-46,737 and 133,388-133,984 in Reference sequence NC_063383) as previously described (Clark et al., 2023) using Q5 Hot-Start High-Fidelity Polymerase (New England Biolabs, Catalog number M0493S) with the following primers: 1-F 5’-ACAGGGTTAACACCTTTCCAATA-3’ + 1-R 5’-AATCTCCAGAACCAGCATCAC-3’ and 2-F 5’ TACAGTTGAACGACTGCG 3’ (Ghate et al., 2023) + 2-R 5’-CTCTCTTGCTTCTTCGTCATAG-3’. The isolated virus was propagated by infection of VeroE6 cells at a Multiplicity of Infection (MOI) of 0.2 and culturing until CPE was observed. Cells were scraped and subjected to 3 cycles of freeze/thaw with vortexing, then clarified by centrifugation, aliquoted, and stored at −80°C for experiments.

MPXV viral stock was titered by plaque assay on VeroE6 cells. Cells were plated in 12-well plates one day before infection. The virus was tenfold serially diluted and incubated on cells with rocking for 1h, then removed and replaced with 0.8% methylcellulose in MEM (Thermo Fisher Scientific, Catalog number:12492013) supplemented with 2% FBS and 1x Penicillin/Streptomycin (Thermo Fisher Scientific, Catalog number: 15140122). After three days of cell growth at 37°C and 5% CO_2_, monolayers were fixed in 4% formaldehyde and stained with crystal violet to visualize plaques.

MPXV was heat-inactivated at 65°C for 40 min in a thermocycler. Before removal to BSL2, complete inactivation was confirmed by plaque assay and by culturing 10% of the material on VeroE6 cells for > 1 week, followed by blind passaging. Cells were monitored for CPE, and media was collected to track copies of the viral genome using the above primers to assay for an increase in viral copies.

HSV-1 from a positive patient sample was isolated on BHK-21 cells (ATCC, CCL-10) by applying serial dilutions of the sample made in RPMI (Thermo Fisher Scientific, Catalog number: 11875093) + 20mM HEPES to monolayers of cells with rocking for 1h, then adding DMEM + 10% heat-inactivated FBS + 1x Antibiotic/Antimycotic. Supernatants were harvested when CPE spread throughout. As described above, the virus was titered by a plaque assay on VeroE6 cells, except that the overlay was 0.43% agarose in DMEM supplemented with 2% FBS and 1x Penicillin/Streptomycin. HSV-1 was heat-inactivated as described for MPXV, and inactivation was confirmed by plaque assay.

MPXV and HSV-I were isolated from consented patient samples under UC San Diego IRB #160524. All work with infectious MPXV was conducted under Biosafety Level (BSL) 3 conditions following guidelines approved by the Institutional Biosafety Committee.

Cowpox virus (CPXV), strain Germany_1998_2, was grown in confluent flasks of BSC-40 cells. (ATCC, CRL-2761). Cells were infected at an MOI of 0.1 and incubated for 48-72 hours before harvesting. Cells were scraped, pelleted at ∼1000 x g, the supernatant removed, and the cell pellet resuspended in RPMI + 2% FBS before undergoing 3x freeze/thaws and sonication before aliquoting. The virus was titered using serial dilutions in 6-well plates of confluent BSC-40 cells. After incubation for 48 hours, plaques were visualized with a crystal violet stain and counted. The resulting titer was 1.1 x 10^8^ PFU/ml. Virus aliquots were then inactivated with gamma irradiation (4.4 x 10^6^ rads) while frozen on dry ice. The CPXV work was conducted at the BSL2+ facility, and the CDC provided samples.

### Enzyme-Linked Immunosorbent Assay (ELISA)

The ELISA assay was performed as described previously (Ray et al., 2024). Briefly, the inactivated viruses (MPXV, HSV-1, and CPXV) at the indicated PFU/mL were diluted in the carbonate-bicarbonate (pH 9.4) buffer (ThermoFisher Scientific Catalogue: 28382). For coating the 96-well microtiter plates (Sarstedt, Catalogue: 82.1581.100), a 100 μL volume of these diluted viruses was added directly to each well, sealed with adhesive strips, and incubated overnight at 4°C. The contents of the plates were discarded the next day, and the wells were washed with 200 μL PBS buffer. Following this, 200 µL of blocking buffer (1x PBS with 1% BSA) was added to each well, covered with adhesive strips, and incubated at room temperature for two hours to block the remaining well surface unoccupied by the antibodies. This step improves the assay’s sensitivity by reducing the background signal and increasing the signal-to-noise ratio. The blocking buffer was discarded, and 100 μl of the anti-A29 monoclonal antibody (mAb 69-126-3-7) (Laura J. Hughes et al., 2014) was added to each well at 1:3000 dilution in 1x PBS with 1% BSA buffer. The concentration of the antibody stock was 2.228mg/ml. The plates were then covered with adhesive strips and incubated for 90 min at room temperature on the rocker. The samples were discarded, and the wells were washed four times with 200 µL of PBS buffer. Next, 100 µL secondary antibody, Goat anti-mouse-HRP (Thermo Fisher Scientific Catalogue: 31430) at 1:4000 dilution, was added, and the plates were sealed with adhesive strips and incubated at room temperature for one hour on the rocker. The samples were discarded, and the wells were washed four times with 200 µL of PBS buffer. 100 µL of 3,3′,5,5′-Tetramethylbenzidine (TMB) substrate solution (Thermo Fisher Scientific Catalogue: N301) was added to each well, waited for two minutes, and the 100 µL TMB stop solution (Thermo Fisher Scientific Catalogue: N600) was added. The plates were scanned within 15 minutes in the Tecan Multimode microplate reader (Spark®) at 450 nm.

All the assays were conducted in triplicate (n=3) sets at every concentration for statistical significance and Limit of Detection (LOD) calculations. The threshold signal is calculated as an average signal from the negative control (HSV-1) plus three standard deviations. LOD is calculated as the concentration value corresponding to the point where the dilution curve intersects the threshold line.

### PD-IRIS Chip Preparation

The surface of the PD-IRIS chips was activated with oxygen plasma (Plasma Etch, PE-25). Subsequently, a polymer-based coating (MCP-4, Lucidant Polymers) was applied to the activated surface of the chips, which covalently binds amine groups. The chips were immersed in a 1X polymer solution (1% w/v polymer in 20% saturated ammonium sulfate) for 30 min. The chips were then thoroughly rinsed with DI water, dried with nitrogen, and baked in a vacuum oven at 80 °C for 15 min. The chips were stored in a desiccator until microarray spotting.

The capture probes immobilized on the surface were 0.5 mg/mL protein G (Millipore Sigma, Catalog number: 08062) and 0.5 mg/mL streptavidin (Prospec, Catalog Number: Pro-791-b). The protein solutions were spotted in 200 mM sodium phosphate buffer with 0.01% Trehalose, pH 8.0, on the MCP-4 coated chips using an M2-Automation iONE-600 spotter. The probes were then incubated in a high-humidity chamber overnight (18 hours) to allow immobilization. After incubation, any remaining active groups on the polymer surface were blocked using a 50 mM ethanolamine solution in 150 mM Tris-HCl, pH 9.0. The chips were incubated in the blocking solution for 1h at room temperature and then washed with 1X PBS with 0.05% Tween. After rinsing in DI water, the chips were dried with nitrogen before assembly.

### PD-IRIS Prototype

PD-IRIS is a wide-field imaging technique that utilizes epi-illumination, i.e., reflected light microscopy. A novel illumination device with two LEDs (410 nm and 660 nm) provides a simple, cost-effective illumination source for uniformity corrections (Çelebi et al., 2023). This light source, EUCLID (Efficient Uniform Color-Light Integration Device), uses an adjustable hollow cavity that enables uniform light mixing from two different input ports. The uniformly mixed light is then introduced to the optical system after it passes through an adjustable iris. This output is imaged on the back focal plane of the objective lens (Nikon, Super Plan Fluor, 20x, 0.45NA) by two achromatic lenses (AC254-075-A, Thorlabs). The diameter of the iris is set to 4 mm to provide low-NA Koehler illumination to the sample chip surface. The reflected light is collected by the same objective lens, and it is imaged onto a CMOS sensor (Flir, BFS-U3-244S8C-C) using a tube lens (TTL 200-A, Thorlabs). The sensor’s exposure time is set to 16 ms and runs at a speed of 16 frames per second.

PD-IRIS substrates, 60 nm-oxide silicon chips, are functionalized as described in the “PD-IRIS Chip Preparation” section to capture and immobilize the virions. The chip has two laser-cut holes to introduce sample buffer in and out over the active area. The side and top border of the chip channel are built by attaching a glass cover on the chip using a pressure-sensitive adhesive gasket. The gasket has a 1.5 mm rectangular opening in the center, allowing for imaging of the active area during the incubation. The chip is inserted into a custom chip holder with fluidic connections that provide samples to the chip through its laser-cut holes. The custom holder is mounted onto a 3-axis Nanomax stage (MAX312D). The optimal focus is determined by inspecting the silicon-etched marks on the chip, which are adjusted by a differential drive and locked to avoid any drift.

The flow in the channel is governed by a programmable 500 µL syringe pump (Hamilton, PSD4) with an 8-input valve to select between wash and sample channels, and the channels are connected by 0.01” ID tubes. The pump speed is 1500 µL/min and 10 µL/min for wash and sample channels. The wash channel is connected to the chip through the pump valve, whereas the sample channel is directly connected to the chip to decrease the dead volume introduced by the valve. 100 µL viral samples flow back and forth 3 times, corresponding to ∼1-hour incubation.

The prototype is controlled by custom-written Python code. The camera and pump can be addressed simultaneously, enabling acquisition when the pump runs for incubation. At the end of the experiment, the acquired images are processed by another custom-written Python code to detect and count individual virions.

### Limit of Detection Determination for MPXV detection with PD-IRIS

A homogenous assay protocol is employed to optimize the capture efficiency of the chip surface (Seymour et al., 2015). In a homogenous assay, the virions are statically incubated with their corresponding antibody outside the system’s fluidic channel (i.e., in an Eppendorf tube), and a molecule that has a high affinity with the antibody is spotted on the chip surface. Then, antibody-decorated virions are flow-incubated over the spotted chip. Different MPXV dilutions are statically incubated for our experiments with the anti-A29 mAb, and protein G is spotted on the chip surface to capture flowing antibody-virus complexes.

For the limit-of-detection (LOD) experiments, three different virus samples are prepared at 5×10^4^ PFU/mL, 1×10^4^ PFU/mL, and 1×10^3^ PFU/mL, which also contains anti-A29 mAb at 2.5 ng/mL. To prepare homogeneous virus-mAb mixtures, the stock mAb solution (at 2.228 mg/mL) is diluted to 0.01 µg/mL in 1x PBS, filtered with 0.02 µm Whatman Anotop filter. The stock MPXV sample (4.6×10^6^ PFU/mL) is diluted in 1x PBS solution to 1×10^5^ PFU/mL, 2×10^4^ PFU/mL, and 2×10^3^ PFU/mL. Experimental samples are prepared by mixing 50 µL diluted MPXV sample with 25 µL of diluted antibody solution and 25 µL of filtered 1x PBS to make a homogenous sample of 100 µL. Then, the prepared sample is left for static incubation for 5 minutes.

The flow incubation protocol consists of three steps. First, filtered 1x PBS solution is flowed over the protein G spotted chip twice at 1500 µL/min as an initial wash step for 20 seconds per wash. Then, the waste is replaced by a homogenous sample, and the syringe pulls and pushes the virus solution three times consecutively over the chip at 10 µL/min, corresponding to 65 minutes of flow incubation. After the incubation, the chip is washed twice at the same speed to remove non-specific bindings. During the flow incubation of the homogenous sample, the camera acquires spot images constantly at 16 frames per second. The acquired images are averaged every 30 seconds to create one data point in a real-time binding curve. After the ∼1 hour incubation and washing, the surface-bound particles are counted for both the protein G spots and negative spots and divided by the spot area to express the bound particle density as particles/mm^2^.

The detection threshold is calculated from the HSV-1 homogenous assay, in which the HSV-1 sample is mixed with anti-A29 mAb and flowed over the PD-IRIS chip using the same steps described for the MPXV assay. The detection threshold is calculated as the mean particle density on the protein G spots (n=6) plus three standard deviations. LOD for MPXV detection is determined by linearly extrapolating the two lowest concentration data points and finding the concentration value at the point where the line intersects the threshold signal line.

### Specificity Experiments using HSV-1 and CPXV

The specificity experiments are also performed using the homogenous assay protocol described in the previous section. The stock virion samples (9.67×10^7^ PFU/mL for HSV-1 and 1×10^7^ PFU/mL for CPXV) are diluted in filtered 1x PBS down to 2×10^4^ PFU/mL. Then, 50µL of diluted samples are mixed with 25 µL of monoclonal antibody solution and 25 µL of filtered 1x PBS to top the volume of the homogenous sample up to 100 µL. The final viral concentration of the homogenous samples is 1×10^4^ PFU/mL.

The same incubation protocol is followed for specificity experiments. The protein G spotted chip surface is washed twice at 1500 µL/min with filtered 1x PBS. Then, homogenous virus-mAb mixtures are incubated for 65 minutes at 10 µL/min. Finally, the incubation is followed by two wash steps. The data points for the flow incubation are calculated by averaging the accumulated images every 30 seconds.

### Proof-of-Concept Experiments

Anti-IgG-coated 80 nm gold nanospheres (Nanopartz) are used and immobilized on the IgG-spotted 60 nm oxide PD-IRIS chip for the proof-of-concept experiment. The chip surface is washed with PBS before and after flowing the GNS-mAb sample at 50 µl/min for 10 min with the syringe pump. The concentration of the GNS is 10^7^ particles/mL (∼10fM), which is diluted in 1x PBS.

### Particle Detection and Tracking Algorithm

In PD-IRIS setup, the camera acquires images constantly before, after, and during the flow incubation. The captured 8-bit images are added onto a float-64 array. That array is averaged every 30 seconds, creating one averaged image, and the resulting image is passed to the particle detection and tracking algorithm to quantify the bound particles to the surface after some pre-processing steps. In the pre-processing part, the first step is to extract spot locations from their corresponding circular etched regions. After the spots are located for every frame, the background levels of different color channels, corresponding to reflected light from the spots, are mapped to the same readout value by dividing each channel by its mod. After the peak values of different color channels are set to the same value, the fixed pattern noise (or pixel-to-pixel variation) is removed by applying the look-up table to the normalized image.

The signal extraction from pre-processed Bayer images has two different methods for experiments. In the proof-of-concept experiment, an individual gold nanosphere image is convolved with a variation filter, resulting in the following constructed signal *I_signal_*_,l,_

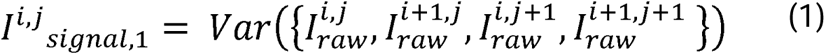

Where superscripts *i* and *j* represent the pixels of the constructed signal and the raw image. The second operation involves convoluting the raw image with two different 2×2 matrices 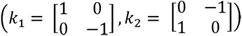 in parallel. The element-wise square of the resulting matrices was summed to construct signal *I_signal_*_,2_, which can be written as,

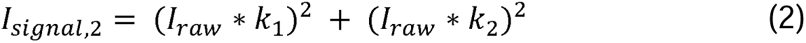

Once the signal is constructed, the particles must be detected and counted for every frame. To do this, the signal is correlated with a 32 x 32 Gaussian function whose width is dictated by the optical resolution (λ/2*NA*). Then, an arbitrary threshold is applied to segment the highlighted features in the correlation result. All of the white regions in the resulting segmented image are detected by OpenCv “*findContours*” function (Itseez, 2015), and the particle candidate locations are identified by filtering the detected contours given the optical properties of the setup (i.e., diffraction-limited size of the particle, circularity of the contour, etc.). Those locations are passed into the tracking algorithm.

The particle tracking algorithm is based on SPANDEX, developed by Sevenler et al. (Sevenler et al., 2019). It accepts the candidate particle locations and tracks each particle’s appearance within a pre-defined region in the time stack using Trackpy (Allan et al., 2023). The tracking enables kinetic assay screening and eliminates the non-specifically bound or falsely detected particles, given that such artifacts cannot appear in the same location for multiple frames. All detection and tracking codes are written in Python, which facilitates the POC applicability of the design.

## RESULTS

### Multi-Spectral Pixel-Diversity IRIS

Pixel-diversity IRIS improves upon the existing Single Particle Interferometric Reflective Imaging Sensor (SP-IRIS). SP-IRIS is a well-established technology that detects and characterizes individual biological particles such as viruses, bacteria, and exosomes in a label-free fashion, as well as biomarkers with the use of nanoparticle labels (Seymour et al., 2023b; Xia et al., 2023; Zaraee et al., 2020). The method utilizes a silicon-silicon dioxide chip on which a microarray is spotted to screen multiple target particles. The captured particles are excited with a precise light wavelength determined by the target particle and the oxide thickness in a common-path interferometric system. The optical response of the particle, *E_s_*, and reflected light from the chip surface, *E_r_*, are imaged on a CMOS sensor on which they interfere. In the presence of small biological particles (*r* ≪ λ), the interferometric cross term dominates the scattering intensity. Thus, the signal contrast is defined as,

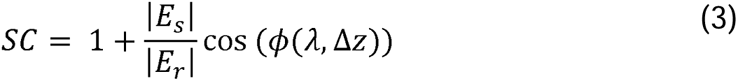

As Eq.3 suggests, the signal relies not on scattering intensity but its amplitude (*SC* ∝ *r*^3^) in SP-IRIS. Although there is a tremendous signal enhancement, distinguishing target particles from the background becomes challenging with wide-field, low-magnification objective lenses. Therefore, the focus dependence of the phase angle (Eq.3) is exploited in SP-IRIS by taking multiple defocus acquisitions, and the signal is constructed by calculating the pixel-wise variance of those acquisitions. In this case, the signal-to-noise ratio (SNR) depends on the number of captured images. This creates a tradeoff between temporal resolution and SNR. Another disadvantage of z-acquisitions is that they require repeatable, high-resolution scanning optics, like a piezo scanner, significantly increasing the cost.

In multi-spectral PD-IRIS, the need for the high-resolution scanning optics is eliminated. The wavelength dependence of the phase angle in the interferometric cross-term (Eq.3) is exploited to increase the faint signal contrast of a small particle. As a result, the sample can be screened at higher speeds, the data takes up less space on the computer and can be processed with more efficient algorithms. This makes the configuration more cost-efficient and rapid for point-of-care testing. Detection at a single snapshot consists of two steps: (i) illuminating the particles with multiple colors simultaneously and (ii) recording the response with a conventional color camera, resulting in checkerboard patterns in the camera readout in the presence of particles.

The differences between signal construction methods are demonstrated in Figure 2. Anti IgG-coated gold nanospheres are captured on the IgG spots to generate SP- and PD-IRIS signals. In SP-IRIS, 31 defocus images are taken across the optimal focal plane with 20x magnification using 530 nm dominant LEDs and stored in a 3D cube. Then, pixel-wise variations are calculated along the z-dimension. In this method, some spot features, like edges, may generate false signals, which increase the background level. Figure 2b demonstrates a PD-IRIS measurement using the same particles in Figure 2a. The particles are simultaneously excited under RGB light (460 nm, 523 nm, and 630 nm). The standard deviation within each superpixel is calculated to generate the processed image. With this new detection technique, PD-IRIS yields an improved SNR, and some image artifacts due to defocus, like spot edges, can be removed.

**Figure 2:**
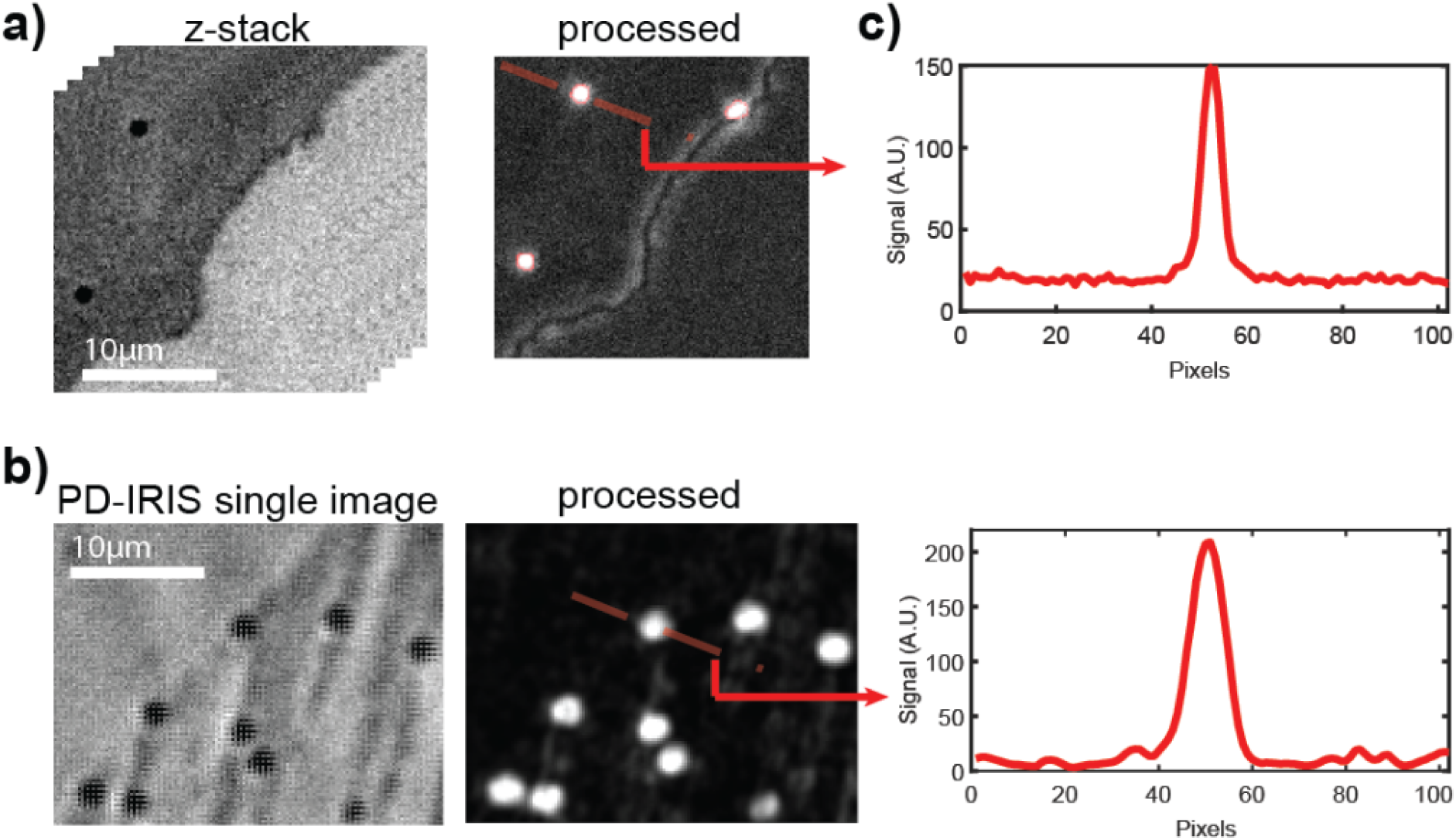
Comparison of SP-IRIS (a) and PD-IRIS (b) modalities. The signal along the cross-section of the analyzed particle for both techniques is given in (c).

The optical signature of the particle is encoded in the high-frequency components of the captured PD-IRIS image. Other high-frequency features/noise must be removed to recover this signal with high SNR. One possible noise originates from the offset of the color channels on the spots. The reflected light from the spots depends on the spot thickness and the illumination wavelength. Since the thicknesses can vary, the value distribution of the spot pixels would be different in every color channel when the LED intensities are not adjusted properly, resulting in a significant background level. This effect is eliminated by detecting every spot in the field of view and correcting the pixel readings with respect to the channel’s mod value. To facilitate this post-processing step, a circular array is etched on the active area of the Si-SiO_2_ chip, indicating the location of the spots. Due to the huge contrast difference, the spots can be detected by simple thresholding, and channels are normalized once an individual spot is cropped from the FOV.

Multi-spectral illumination’s spatial and spectral non-uniformity is another noise source that would reduce the SNR. Similar to channel offsets, any non-uniformities in the illumination would increase the background level in the constructed PD-IRIS signal. Thanks to our group’s recently developed light source, EUCLID (Çelebi et al., 2023), multiple colors can be spatially mixed with exquisite uniformity and inputted to the imaging optics from one output port. EUCLID’s simplicity, cost, and size make it appealing for a POC setup.

The last two high-frequency features we studied are shot and fixed pattern noise. The shot noise can be reduced by collecting more electrons by averaging sequential frames. However, SNR cannot be improved further by simply averaging more frames because fixed pattern noise or pixel-to-pixel variations (Zapata-Pérez et al., 2020) become the dominating factor. Reading errors due to pixel-to-pixel variations introduce a significant noise source when particle signature is expressed in the sudden changes between the adjacent pixels. Thus, this must be removed for high SNR PD-IRIS images. This issue can be mitigated by using a uniformly illuminated mirror image as a look-up table and correcting every image accordingly.

### Proof-of-concept Experiments with PD-IRIS

To test the optical performances of different objective lenses and determine the optimal focal position for PD-IRIS, we measured the defocus signature of 80 nm GNSs and the immobilized MPXV particles. First, we tested color aberrations introduced by the imaging optics. We imaged the immobilized GNSs with 10x (Nikon Plan Apo λD) and 20x (Nikon Super Plan Flour) objective lenses and compared the focal shifts of different colors (Figure 3a-b). Since the 10x objective was designed to minimize the color dispersion, the focal positions for different wavelengths were maintained. The 20x objective lens, however, yielded a –1.5µm shift for the 530 nm dominant green and a +1 µm shift for the 405 nm dominant blue LEDs. Unlike in a regular wide-field microscope, this focal shift increases the signal contrast difference between different wavelengths and improves PD-IRIS performance.

**Figure 3:**
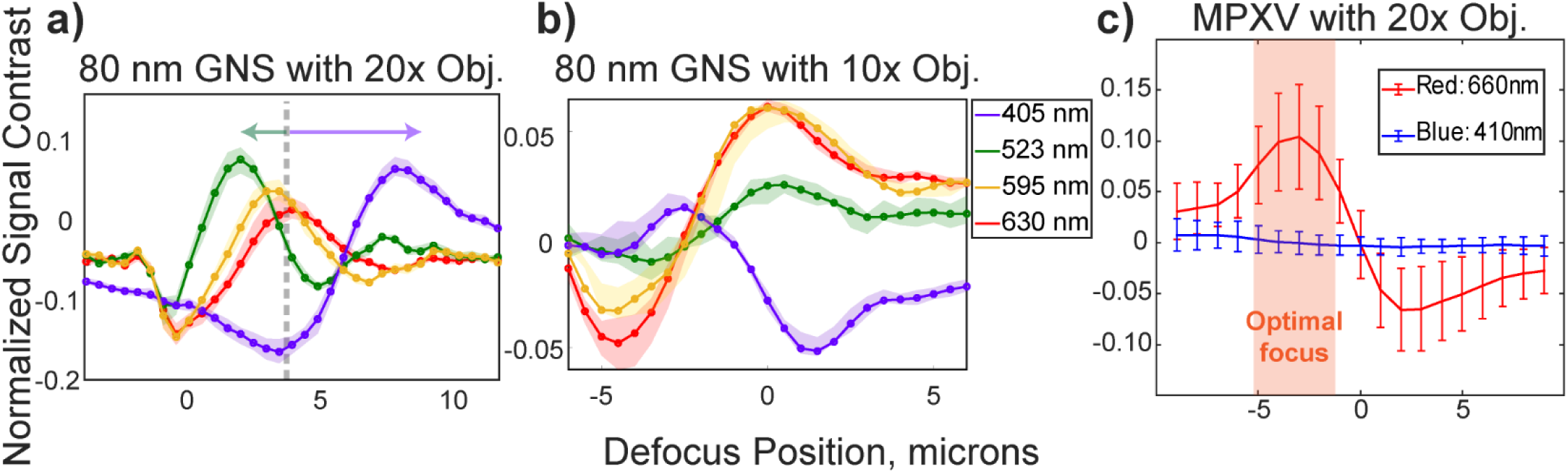
Defocus profiles of 80 nm gold nanospheres (GNS) (a, b) and MPXV particles (c) that are immobilized on silicon chips with a 60 nm SiO_2_ top layer. The measured defocus signals are shifted when the light is collected with 20x S Plan Flour, Nikon objective lens. The arrows indicate the shift due to chromatic aberrations (a). A 10x Plan Apochromatic Lambda D, Nikon objective lens doesn’t yield defocus shifts (b). All defocus profiles have a ∼ 5 µm focus range where the difference between minimum and maximum contrast of different color channels is greater than 1%. The shaded region indicates where the focus is set for MPXV experiments (c).

Finally, we determined the optimal focus location for MPXV particles by comparing its defocus profile at 410 nm and 660 nm. The selected wavelengths were also used for all virus detection experiments. Selecting two well-separated wavelengths was to reduce the crosstalk between color channels. As indicated in Figure 3c, there is ∼5 µm optimal defocus region. This focus region can be found manually by using differential drivers instead of piezo stages, which would be a one-time adjustment for the user and reduce the cost significantly.

### MPXV Detection Experiments with PD-IRIS

Next, we determined our PD-IRIS assay’s sensitivity and specificity for MPXV detection and compared our sensor data with ELISA results. The experiments are performed as described in the *Materials and Methods* section. We opted for a homogenous assay procedure in the experimental design due to its advantages in increasing the assay sensitivity and decreasing the assay time. The virus solutions are first incubated with an anti-A29 monoclonal antibody to allow for virus-mAb complex formation in this assay type. Virus experiments are performed in the prototype PD-IRIS setup (Figure 4a). The PD-IRIS chips are printed with a microarray consisting of protein G (positive), which has a higher affinity for mouse monoclonal antibody than protein A (Fishman and Berg, 2019), and streptavidin spots as the negative control (Figure 4c). The fluidic channel for the flow incubation is built in three layers (Figure 4b). A cover glass encapsulates the buffer solution when assembled to the functionalized PD-IRIS chip via a pressure adhesive tape.

**Figure 4:**
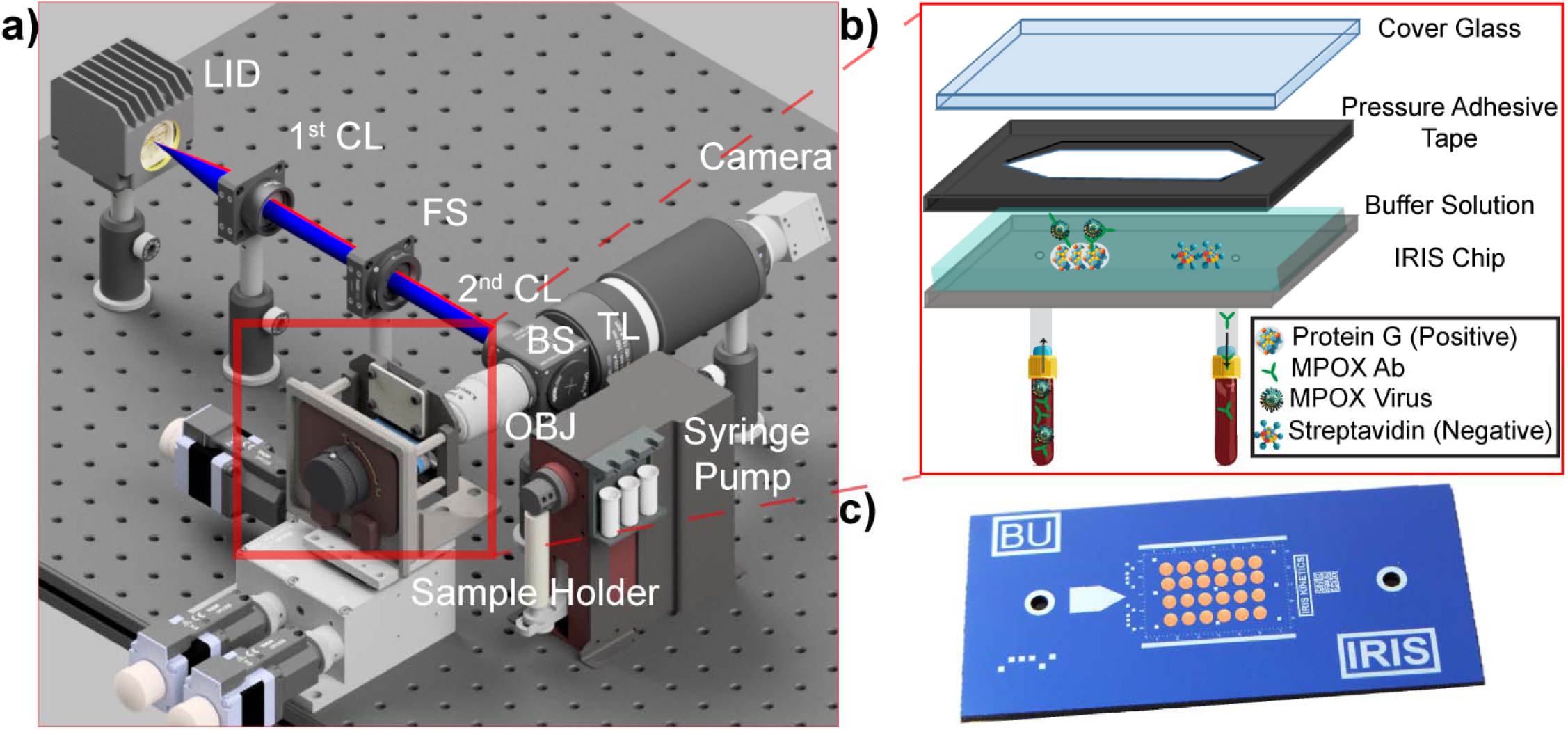
Prototype of PD-IRIS (a). Uniformly mixed light at two distinct wavelengths excites the sample in the Koehler configuration. The reflected and scattered light response is collected by the same objective lens and imaged onto a conventional color CMOS sensor. The chip and glass cover are assembled using pressure adhesive tape, creating a fluidic channel for the sample incubation (b). An image of the PD-IRIS chip with microarray spots printed (c).

First, we optimized the antibody concentration in homogeneous virus detection experiments to maximize capture efficiency. Unlike heterogenous assays, where the antibodies and virions are incubated sequentially on the active area of the chip, antibody-covered virions are introduced into the channel in a homogenous assay. In this assay format, since the antibodies are added in excess of the virus particles, the antibody concentration needs to be adjusted so that free antibodies in the solution do not saturate the protein G spots. An optimized homogenous assay is more efficient than a heterogenous assay in terms of capture efficiency (Seymour et al., 2021), and sample-to-result time is much shorter, considering it only involves one flow incubation. We mixed the virus samples (1×10^5^ PFU/mL) for the antibody optimization experiments with three different antibody concentrations (2.5 μg/mL, 2.5 ng/mL, 0.25 ng/mL). Given those concentrations, the antibody-to-viral particle ratio is determined and provided in Supplementary Table 1. The 2.5 ng/mL mAb concentration was selected as it yielded the highest signal for detected viral particles (Supplementary Figure 1).

PD-IRIS utilizes a 1’x0.5’ chip format with a 5 x 25 circular etched array to indicate the spot positions. For our preliminary results with the MPXV, we consider two FOVs covering six protein G (positive) and four streptavidin (negative) spots. The number of captured virions on the positive and negative spots within an FOV is averaged separately to eliminate the spot-to-spot variations due to printing imperfections. First, the Limit of Detection (LOD) is estimated, the results of which are shown in Figure 5a. We conducted experiments with three serial dilutions of MPXV (5 ×10^4^, 1 x 10^4^, and 1 x 10^3^ PFU/mL) and a blank sample (HSV-1 at 10^4^ PFU/mL) on separate chips. PD-IRIS could detect viral particles at 10^3^ PFU/mL, showing a signal well above the threshold. PD-IRIS achieved a calculated LOD of ∼200 PFU/mL from the extrapolated curve, outperforming ELISA by almost one order of magnitude (LOD ∼1800 PFU/mL) (Figure 5b). It should be noted that in the case of ELISA, although we are coating the plates with inactivated MPXV, there may be free A29 protein in the solution that can contribute to the signal. Whereas, with the PD-IRIS system, we only detected the whole MPXV, not the free A29 protein. For MPXV (Orthopoxvirus), the Viral Particle (VP)-to-PFU ratio is 10 (Americo et al., 2017). Therefore, the PD-IRIS’s LOD - 200 PFU/mL corresponds to 2000 VP/mL. Considering 1 mole is 6.022 ×10^23^ (*N*_A_ = Avogadro Constant) viral particles, the calculated MPXV LOD translates into 2×10^6^/ *N*_A_ = 3.3 x 10^-18^ mol/L (∼3.3 attomolar).

**Figure 5:**
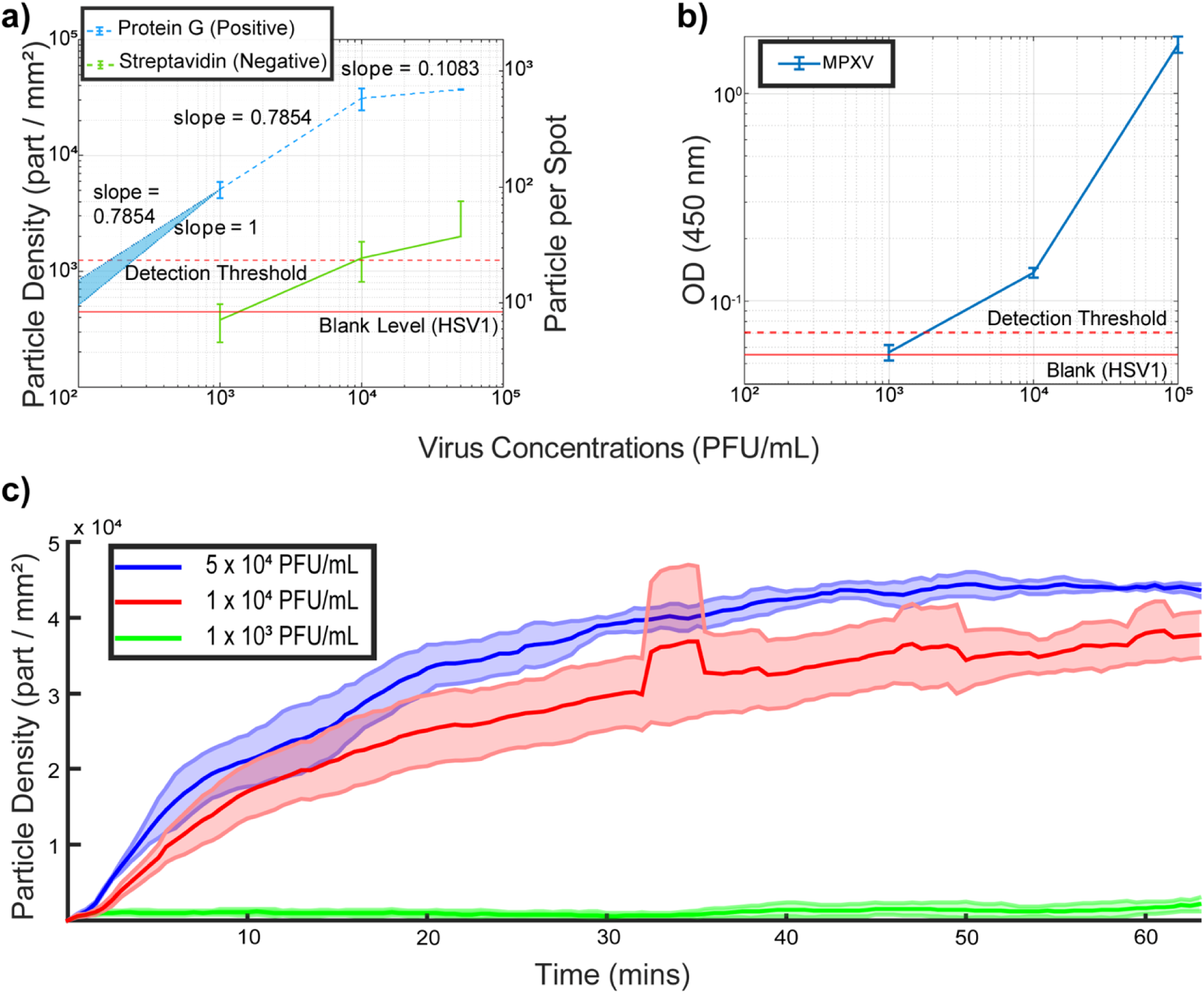
Estimating the sensitivity of PD-IRIS (a) and ELISA (b) for MPXV detection. The signal obtained from the distractor viruses (HSV-1) was used to determine the limit-of-detection (LOD) level, which is shown as a red dashed line (Mean particle density plus three standard deviations). For PD-IRIS measurements (a), two different FOVs, including three protein G and two streptavidin spots, are analyzed to extract the data points. Once the mean particle density and total detected particles are calculated for each FOV, the error bars are created by calculating the mean and standard deviation of those two FOVs. The error bars for ELISA (c) are calculated from three OD measurements. The LOD curve was extrapolated according to the slope between the least two concentrations of the virus dilutions in PD-IRIS data. The real-time MPXV measurements of PD-IRIS are shown in (c) for three different virus concentrations over the course of the experiment. For each real-time experiment, three different protein G spots are imaged. The solid line represents the mean of those three spots, and the shaded area represents their standard deviations.

We also perform kinetic binding measurements with PD-IRIS by real-time data acquisition (Figure 5c). Images are constantly collected with the camera during the incubation at 15 Frames per second (FPS). (A movie and the dynamic graph created from acquired images are shown in Supplementary Video 1, demonstrating real-time virus binding to a protein G (positive) and streptavidin (negative) spot. Obtained images are averaged every 30 seconds to create a data point in Figure 5c. With the help of kinetic measurements, we screen the incubation and validate that it is done without any experimental artifacts such as air bubbles. Real-time screening also enables us to identify all particles, record their interactions with the chip surface, and distinguish between specific and nonspecific bindings. We incubated the virus samples for over 1 hour and monitored the binding as it reached saturation.

Finally, the specificity of the PD-IRIS MPXV assay is characterized using distractor viruses. For this purpose, two homogenous assays, including the Cowpox virus (CPXV), another species of orthopoxvirus genus, and Herpes Simplex Virus (HSV-1), are prepared. The skin lesions typical of Mpox are similar in clinical presentation to those of CPXV and HSV1 infections. Therefore, for proper clinical management, an ideal POC assay should be able to identify Mpox and distinguish it from these prevalent infections specifically. The same incubation protocol used for MPXV is followed. Inactivated CPXV and HSV-1 at 10^4^ PFU/mL are incubated with the A29 mAb at 2.5 ng/mL. After incubation, the tests are loaded to the PD-IRIS sensor, as described previously, and the particles on Protein G spots are counted to obtain the signal. A low background signal level is detected at the protein G and Streptavidin spots for these distractor viruses, with particle densities below the threshold signal (Figure 6a, Supplementary Figure 2). This indicates that the A29 mAb is specific in capturing only the MPXV on the sensor surface to generate the optical signal. The A29 mAb does not capture CPXV or HSV-1 and, consequently, is not immobilized on the sensor surface to generate signals. The ELISA results corroborate the specificity of the A29 mAb against MPXV compared to CPXV and HSV1 (Figure 6b).

**Figure 6:**
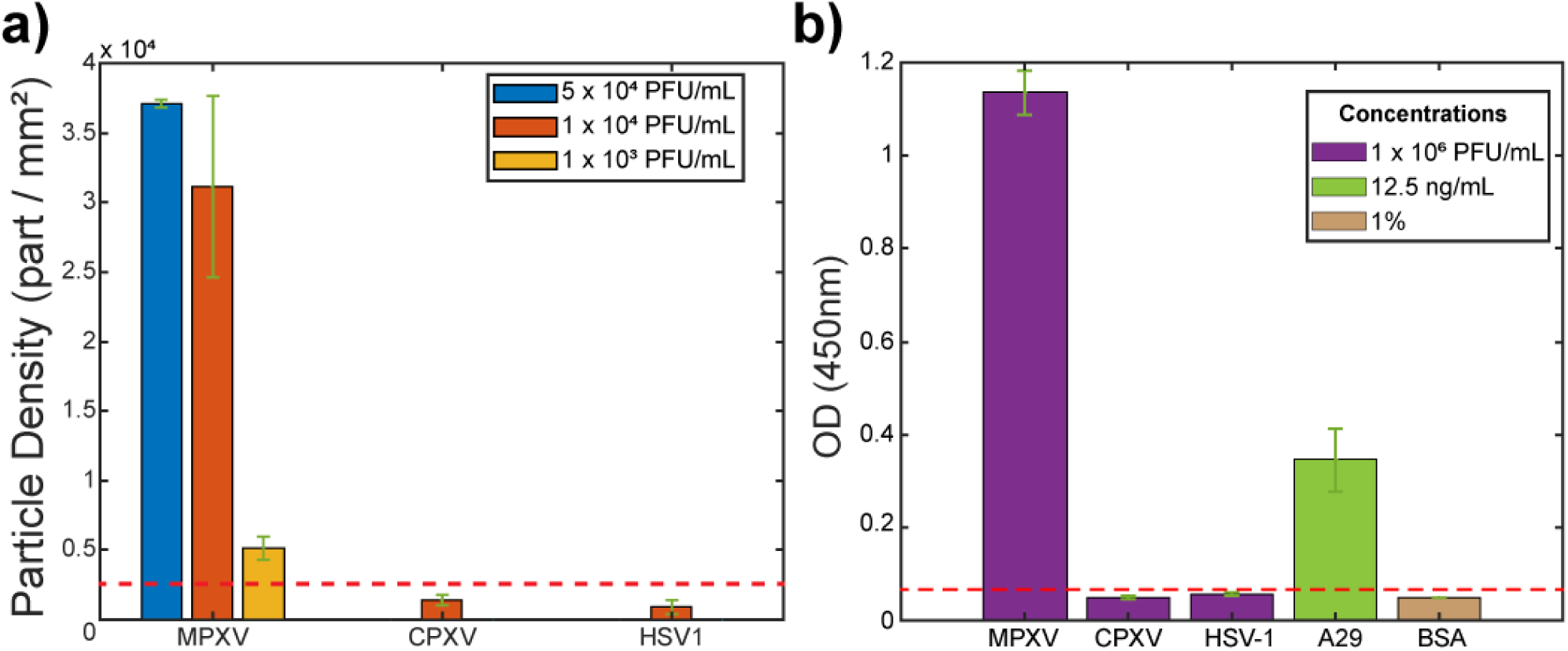
Specificity experiments with PD-IRIS (a) and ELISA (b). The red dotted line represents 3 standard deviations from the mean of the HSV-1 signal for both graphs and corresponds to the detection threshold.

## DISCUSSION

Digital detection, or single molecule counting, is an exciting recent development in diagnostics that provides resolution and sensitivity far beyond ensemble measurements. However, most emerging digital detection techniques, like digital PCR, bead-based single molecule assays (e.g., SiMoA), or digital-ELISA, are based on complicated particle confinement/isolation strategies that increase the cost and complexity of the entire process for POC applications. Pixel Diversity Interferometric Reflectance Imaging Sensor (PD-IRIS) is a technology that utilizes light interferometry from an optically thin film for label-free, high-sensitivity detection of individual nanoparticles (such as viruses or exosomes). As opposed to our previous technology, SP-IRIS, PD-IRIS utilizes multi-parametric particle excitation and special imaging sensors to extract the information encoded in this multi-parametric illumination. In multi-spectral PD-IRIS, the particles are excited with spatially and spectrally uniform light, and a conventional color CMOS sensor is used to extract the encoded particle signal. This novel imaging technique enables particle detection from a single snapshot, eliminating the need for the complicated and time-consuming z-scan process used in SP-IRIS. Moreover, in PD-IRIS, SNR is improved by using a uniform light source (EUCLID) and post-processing steps to decrease the noise from various sources. These advancements offered by PD-IRIS make the system more cost-efficient and faster compared to SP-IRIS, increasing its potential for POC diagnostic applications.

Using PD-IRIS, we demonstrated sensitive and specific detection of MPXV with a calculated LOD of 200 PFU/mL, providing a nine-fold improvement in sensitivity compared to a lab-based alternative test, ELISA. This LOD corresponds to ∼3 attomolar when 1 mol is taken as 6.022×10^23^ (Avogadro Number) viral particles, considering that the viral particle-to-PFU ratio of MPXV solution is 10 (Americo et al., 2017). Moreover, our real-time virus binding experiment shows a significant PD-IRIS signal is achieved in less than 10 min of incubation time for 5 ×10^4^ and 1 x 10^4^ PFU/mL samples. For a workflow similar to the one shown in Figure 1, the total assay time is expected to be about 20 min, including the initial virus sample-antibody incubation step. Thus, our results indicate that PD-IRIS offers a clinically relevant assay platform for detecting early Mpox infections (Paran et al., 2022). Since PD-IRIS detects the virus particles’ surface density (particles/mm^2^), we will establish a calibration curve to correlate the surface density to PFU/mL using Mpox samples with known PFU/mL values. When reporting clinical test results, this information will be incorporated into a conversion algorithm to determine the PFU/mL in the test specimen. Although our MPXV detection experiments were performed in buffer media in the current study, our previous work utilizing SP-IRIS demonstrated that our platform is compatible with complex media such as serum and blood (George G. Daaboul et al., 2014).

We tested MPXV (Clade IIb) in our prototype PD-IRIS system. However, the amino acid sequence of the antigen epitope to which the anti-A29 monoclonal antibody (mAb 69-126-3) binds is conserved between Clade I and II MPXV (Davis et al., 2023). Therefore, our POC assay can most likely be used to diagnose Mpox infection caused by Clade I and II MPXV. Our ELISA data (Supplementary Figures 3 and 4) supports this assumption by demonstrating the detection of A29 protein from Congo basin isolate MPXV-ZAI-96-I-16 (Clade I) using the mAb 69-126-3. (Azzi, 2023).

In our PD-IRIS prototype, we employ a syringe pump to control the flow of the samples. Although the pump can adjust the flow speed precisely, it increases the total cost significantly, which is not desirable for a POC system, and is vulnerable to sample contamination for clinical diagnostic purposes. One alternative to the active fluidic system in our PD-IRIS prototype would be having a passive and disposable cartridge that includes an absorbing pad to conduct the sample flow over the chip (Scherr et al., 2017). Another component that needs to be optimized in our prototype for a POC setup is the 3-axis NanoMax (Thorlabs) stage on which the sample holder is mounted. For an initial prototype and optical characterization of the PD-IRIS setup, we used an expensive 3-dimensional stage and open-loop piezo to tune the focus position precisely. However, it is demonstrated that even with the highest magnification objective lens, 20x, there is ∼ 5 μm of optimal focus position that yields a detectable signal. Thus, a custom stage with a differential driver is enough to adjust the focal distance manually for the experimental acquisition. This will further enable widespread POC applicability of PD-IRIS.

In the recently published article by Song et al. (Song et al., 2024), the authors used an optical biosensor (Fiber Optic Biolayer Interferometer) for label-free detection of Mpox A29 protein at a LOD of 0.62 ng/mL in buffer and 0.77 ng/mL in spiked serum samples. Another study by Zhang et al. (Zhang et al., 2023) used Surface-Enhanced Raman Spectroscopy (SERS) to detect A29 protein at an LOD of 5 ng/mL. However, BLI and SERS are laboratory-based assays that require expensive equipment and trained personnel to run and interpret the tests and are not conducive to POC application (Supplementary Table 2). Also, in our study, we decorated the surface of MPXV virions with an anti-A29 antibody and captured the whole virus on the sensor chips for detection. Thus, compared to the other studies that detected free A29 protein, our assay detected the whole MPXV particles and did not require any sample pre-processing steps, e.g., protein extraction.

The PD-IRIS technology can potentially revolutionize the diagnosis of infectious diseases at the point of care for several reasons: 1) The sensor chips can be stored at room temperature in dry format and do not require refrigeration. 2) The microarray nature of the biosensor substrates allows for readily scalable chip manufacturing, thus lowering the cost per test. 3) Due to its robust signal transduction mechanism, environmental effects such as humidity and temperature do not affect PD-IRIS measurements. 4) Since only virus particles captured by high-affinity probes on the sensor surface are visualized and counted, the PD-IRIS platform is compatible with complex sample matrixes such as saliva, whole blood, serum, or other biofluids. Thus, clinical specimens can be tested directly without requiring preprocessing steps to extract the test material or remove interfering biomolecules. 5) Easy sample preparation and automated data acquisition and analysis make PD-IRIS an easy-to-use platform that simple instructions can operate. 6) Single-particle detection makes the system highly sensitive, enabling detection even at the early stages of the infection.

The recent COVID-19 pandemic has taught us the catastrophic consequences of failing to detect infectious diseases early in their path to curb their global spread. We certainly want to avoid repeating this mistake with the current surge in Mpox infection. Therefore, rapid detection followed by isolation and treatment of the infected patient is our best current arsenal against this rapidly spreading infectious disease. We envision deploying the PD-IRIS as a diagnostic test for Mpox as a partner in this battle. With its high sensitivity and specificity, this test can greatly improve healthcare outcomes by enabling early Mpox detection and substantially reducing the burden of the disease by preventing its spread. Moreover, our PD-IRIS system is a versatile and modular technology. It can be adapted to detect other pathogens with high sensitivity and specificity. This can be achieved by recruiting other high-affinity ligands (aptamers or antibodies) that could specifically bind the pathogens’ surface proteins and capture them on the sensor surface for digital detection.

## CONCLUSION

Digital detection of label-free bioparticles provides sensitivity beyond that of most ensemble measurements, and if made accessible at an affordable price, it can potentially revolutionize disease diagnostics. This work introduced a rapid multi-spectral, label-free detection tool called PD-IRIS. With the PD-IRIS modality, Monkeypox virus detection and enumeration were achieved from single-color images in a robust instrument with no moving parts, bringing us closer to our goal of making this advanced yet easy-to-use disease diagnostic tool available at a low cost. The microfluidic integration reduces sample volume requirements and allows easy clinical specimen collection. The sensitivity of PD-IRIS was also superior to the lab-based diagnostic gold-standard ELISA; thus, it can be deployed to diagnose Monkeypox infection at point-of-care, especially in resource-poor areas. The microarray nature of PD-IRIS substrates also makes it possible to create desired viral diagnostics multiplex panels based on need using specific capture probes.

## Data Availability

All data produced in the present study are available upon reasonable request to the authors

## ACKNOWLEDGEMENTS

The project was partially funded by UCSD Pandemic Response to Emerging Pathogens, Antimicrobial Resistance and Equity (PREPARE) Grant#: 2P30AI036214-29 (PR) and the San Diego Center for AIDS Research (SD CFAR), an NIH-funded program (P30 AI036214) supported by the following NIH Institutes and Centers: NIAID, NCI, NHLBI, NIA, NICHD, NIDA, NIDCR, NIDDK, NIMH, NIMHD, NINR, FIC, and OAR. M.S.Ü. received the National Science Foundation, NSF-TT PFI (2329817) award, which partially funded the research work presented in this study. The findings and conclusions in this report are those of the authors and do not necessarily represent the official position of the CDC.

