## Supplementary Materials for "A Label-free Optical Biosensor-Based Point-of-Care Test for the Rapid Detection of Monkeypox Virus"

[4.] Poxvirus and Rabies Branch, Centers for Disease Control and Prevention, Atlanta, GA 30329, USA

[5.] axiVEND, Winter Garden, FL 34787, USA

[6.] Department of Pathology, University of California, San Diego, CA 92093, USA

[7.] Department of Biomedical Engineering, Boston University, Boston, MA, 02215, USA

*Address correspondence to:

Partha Ray, M. Selim Ünlü

***Supplementary Materials:***

**MPXV A29 protein ELISA**

For ELISA with the MPXV A29 protein, the lyophilized A29 recombinant protein from strain *MPXV-ZAI-96-I-16 (Clade I) was purchased (Arco Biosystems, Catalogue number* A2L-M52H3-100 μg) and dissolved in sterile water to make a stock of 400 μg/mL. Next, A29 protein dilutions at the indicated concentrations were made in the carbonate-bicarbonate (pH 9.4) buffer (ThermoFisher Scientific Catalogue: 28382), and 100 μL of the diluted solutions were added to each well. All the subsequent steps followed were the same as described in the *Materials and Methods* for the inactivated viruses. All the assays were conducted in triplicate (n=3) sets at every concentration for statistical significance and Limit of Detection (LOD) calculations. The threshold signal is calculated as an average signal from the negative control BSA plus three standard deviations. LOD is calculated as the concentration value corresponding to the point where the dilution curve intersects the threshold line.

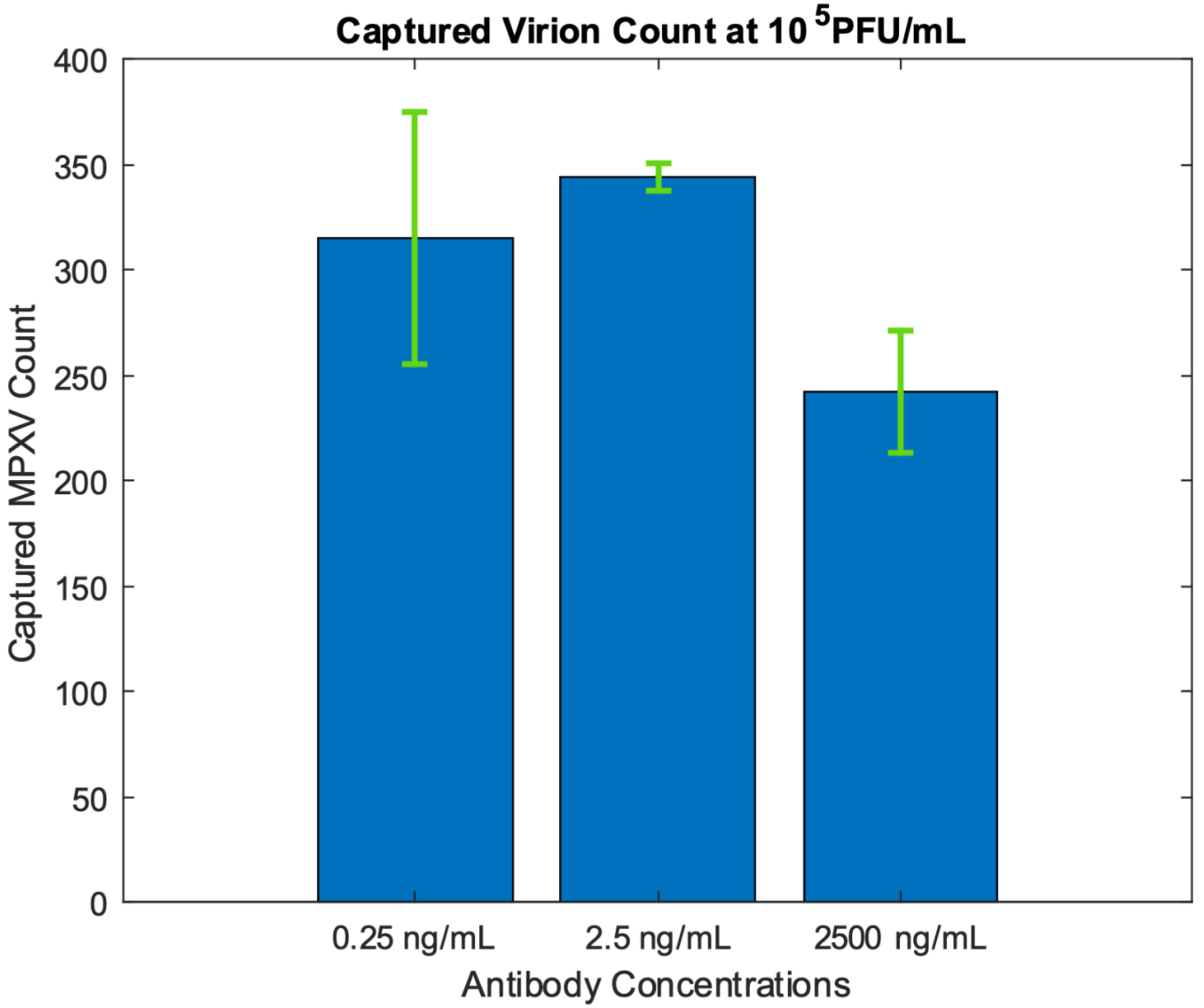


**Supplementary Figure 1:** Captured particle counts of different homogenous assays with varying anti-A29 antibody concentrations. The spot size is the same for all measurements. Three different protein G spots are analyzed for each concentration. The bar graphs represent triplicate sample (n=3) measurements' Mean +/- Standard Deviation (error bars).

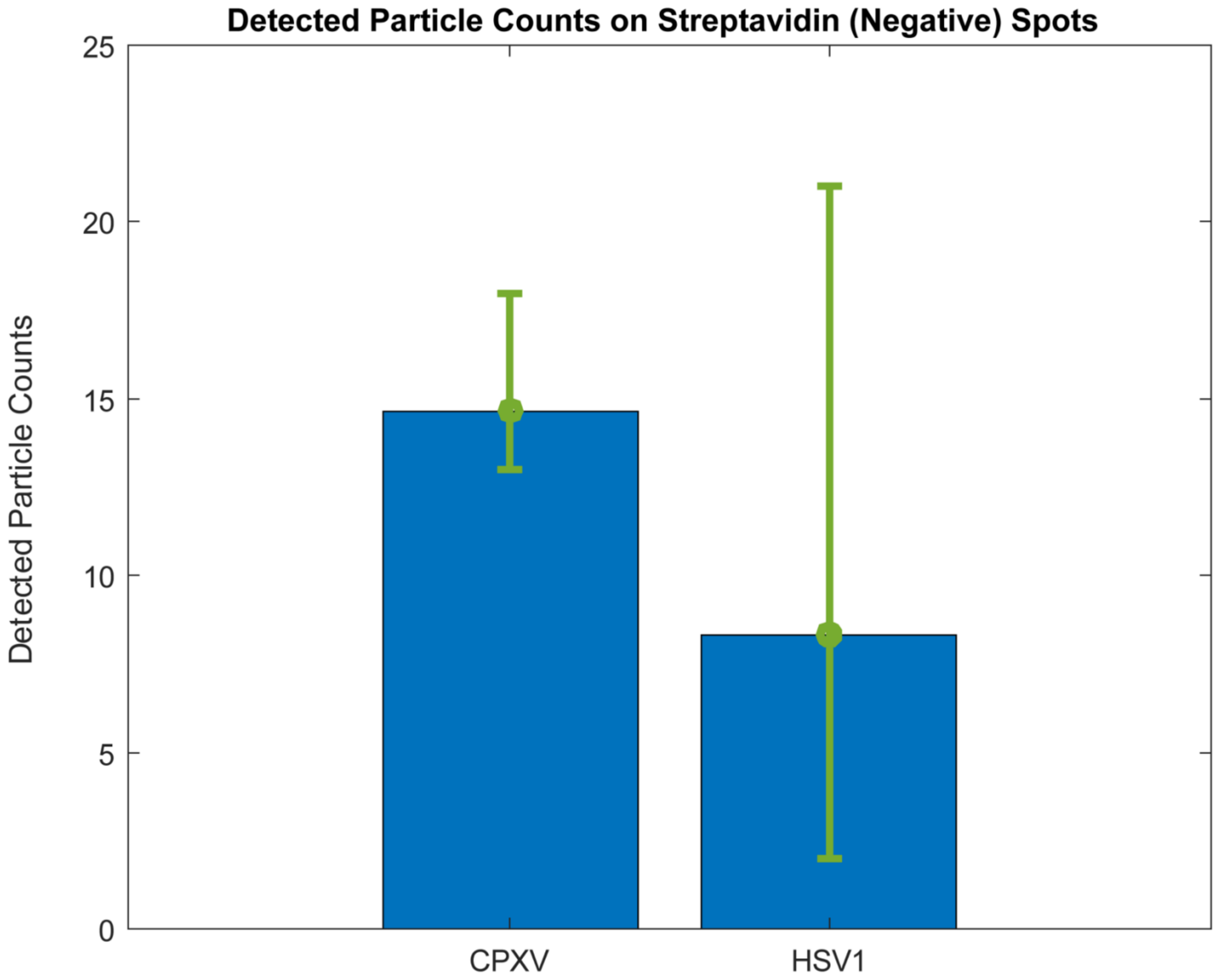


**Supplementary Figure 2:** Detected particle counts on negative control spots for specificity experiments. The spot size is the same for all measurements. Three different streptavidin spots are analyzed for each virus incubation. The bar graphs represent triplicate sample (n=3) measurements' Mean +/- Standard Deviation (error bars).

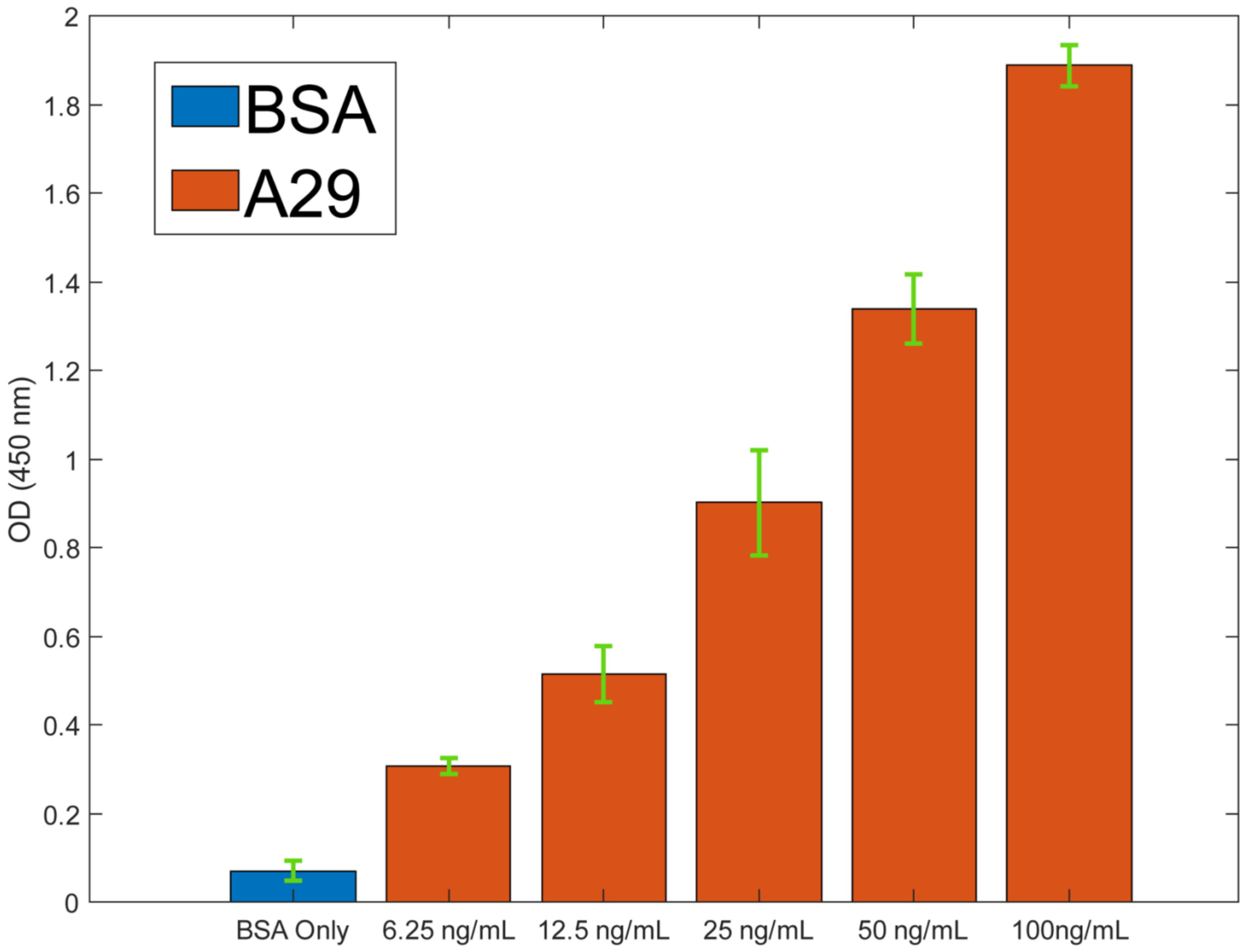


**Supplementary Figure 3:** Results of ELISA experiment to validate the Mpox mAb (mAb 69-126-3) using (Hughes et al., 2014) MPXV A29 protein from strain *MPXV-ZAI-96-I-16 (Clade I)*. The bar graphs show triplicate sample (n=3) measurements' Mean with error bars representing Standard Deviation.

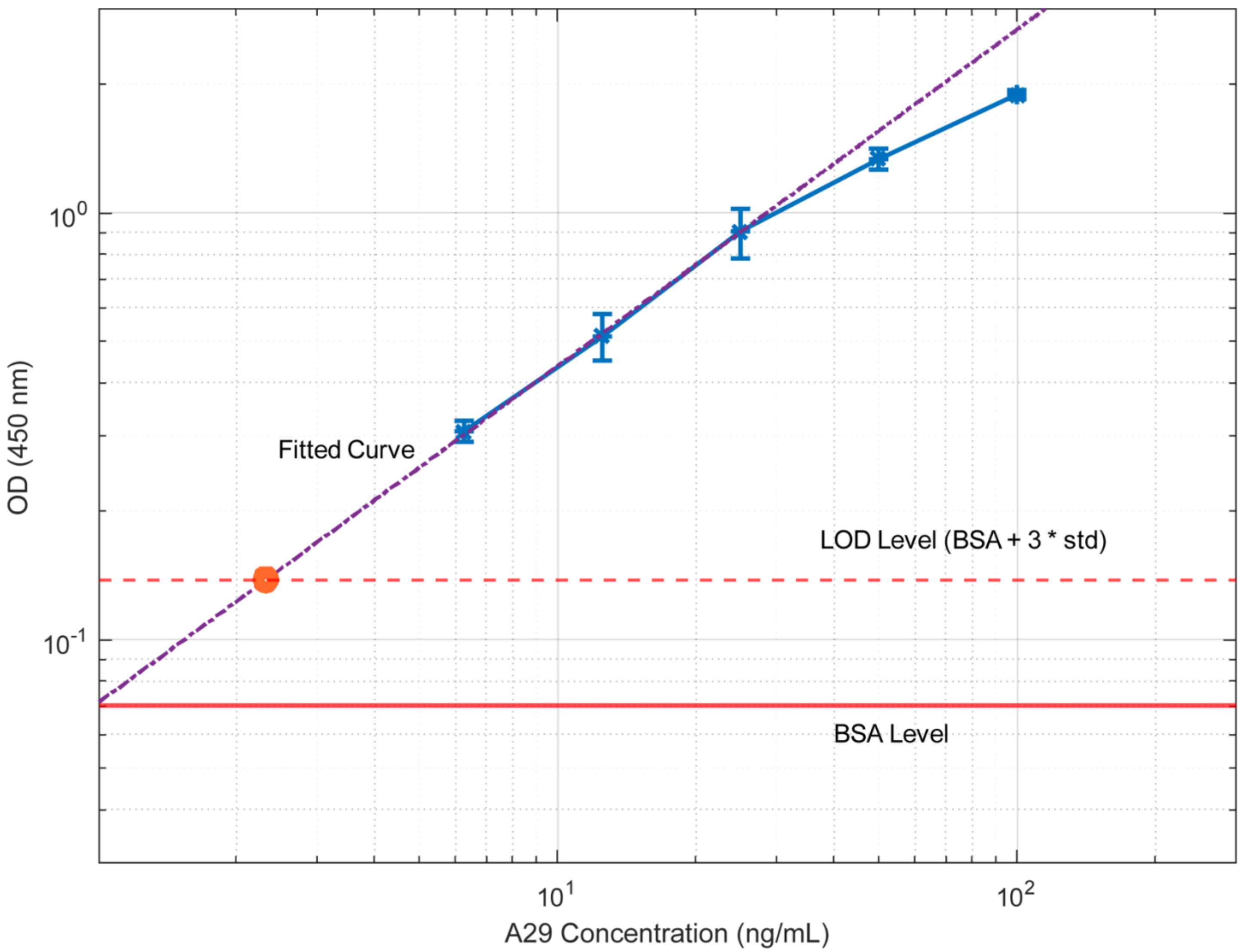


**Supplementary Figure 4**: ELISA LOD curve for A29 protein antigen detection using Mpox mAb 69-126-3. The graph is the mean +/− SD of OD450 values from triplicate samples. The best-fit line was calculated using log-log transformed values. The calculated LOD is ~2.32 ng/mL.

| Antibody / Virus particle Ratio at $10^5$ PFU/mL | Antibody Concentration (ng/mL ) |
|---|---|
| ~8300000:1 | 2500 |
| ~8300:1 | 2.5 |
| ~830:1 | 0.25 |

**Supplementary Table 1:** Antibody to Virus particle ratio calculations. Given that the molecular weight of the anti-A29 antibody is 181 kDa and the viral particle-to-PFU ratio is ~10, the number of antibody molecules in a homogenous assay is calculated from its concentration in (ng/mL)

| Target | LODs in Buffer (Molarity) | Time (mins) | Approaches | POC applicability | Refs. |
|---|---|---|---|---|---|
| A29 | 0.35 nM<br>(5 ng/mL) | 5 | Surface Enhanced Raman Spectroscopy | No | (Zhang et al., 2023) |
| A29 | 43.06 pM<br>(0.62 ng/mL) | 14 | Biolayer Interferometry | No | (Song et al., 2024) |
| **MPXV (Whole Virus)** | **3.3 aM** | **20** | **PD-IRIS** | **Yes** | **This work** |

**Supplementary Table 2:** Comparator table of Mpox detection assays utilizing MPXV A29 antibodies.

**Supplementary Video:** A movie and the dynamic graph created from acquired images demonstrating real-time MPXV binding to a Protein G (positive) and Streptavidin (negative) spot.
